# Plasma glycome reflects low back pain progression and is not predictive of acute-to-chronic pain transition

**DOI:** 10.1101/2024.11.12.24314768

**Authors:** Elizaveta E. Elgaeva, Irena Trbojević-Akmačić, Ivo Ugrina, Marija Vilaj, Anita Slana, Tea Pribic, Yakov A. Tsepilov, Lennart C. Karssen, Yurii S. Aulchenko, Concetta Dagostino, Massimo Allegri, Andrea Skelin, Dragan Primorac, Klaas Buyse, Jan Van Zundert, Gordan Lauc, Frances M.K. Williams, Maxim B. Freidin

## Abstract

**Aims:** To investigate if the plasma N-glycome would provide a biomarker of acute-to-chronic low back pain (LBP) transition.

**Patients & Methods:** A prospective longitudinal sample of n = 1,114 individuals enrolled at the first ever episode of LBP and followed-up for three months. Total plasma N-glycome was measured and compared at the baseline and follow-up.

**Results:** Initial pro-inflammatory patterns of plasma N-glycome change to normal levels between baseline and follow-up in those who resolved from LBP, but not in those who remained at pain after three months. Baseline levels of N-glycans were not predictive of the risk of chronic LBP at follow-up.

**Conclusion:** Total plasma N-glycan levels may reflect ongoing inflammation that is aberrant and promotes ongoing pain and chronicity of symptoms.

## Introduction

Low back pain (LBP) is a common debilitating condition which leads the list of disorders causing disability in much of the world [1]. For the majority of people, the symptoms of LBP are transient; however about 10%-15% develop chronic LBP which is usually defined as LBP lasting for more than three months [2]. It leads to work absenteeism, job loss and long-term disability. In Europe, the costs of chronic LBP are estimated to amount to 1-2% of GDP [3-6]. Risk factors for chronic LBP include age, female sex and raised body mass index, but their relative contribution varies by population [7].

One of the most pressing issues in the field of LBP is our current inability to predict which patients with acute LBP will go on to develop chronic LBP which may be accompanied by disability. The early identification of high-risk patients and stratification into the most suitable treatment groups have been shown to be effective; and the huge economic burden of LBP could be addressed via faster direction of patients to more effective targeted intervention [8]. Several screening tools are used in primary care to stratify LBP patients into targeted treatment subgroups. These screening tools comprise self-report questionnaires that consider physical and psychological characteristics, including pain intensity, functional impairment, catastrophising, anxiety and depression, and predict the risk of persistent pain, disability, recovery, and sick leave duration. The STarT Back Tool (SBT) [9] is one of the most widely used tools of this kind; it allocates patients into low risk group requiring primary care, medium risk group requiring physiotherapy, and high risk group benefiting from a combination of physical and cognitive treatment [10]. SBT and similar tools have been shown to accurately predict the risk of disability and job absenteeism, but their prediction of the development of chronic back pain is rather poor, at 60-70% correct allocation [11].

We hypothesised that combining such a screening tool with serum biomarkers provides a way to improve stratification and prediction of chronic LBP risk. The plasma glycome describes the entire composition of oligosaccharides called glycans attached covalently to proteins and lipids, most commonly to a nitrogen atom of asparagine (N-glycans) or oxygen atom of serine or threonine (O-glycans). Many inflammatory and immune-mediated diseases, as well as some cancers, are characterised by specific changes in the glycome composition [12-15]. Our own studies provided evidence on association between total plasma and immunoglobulin G (IgG) N-glycome with chronic LBP. We identified that higher risk of chronic LBP is associated with the lack of core fucosylation of IgG [16]. Given that core fucosylation of IgG is a “safety switch” for antybody dependent cell mediated cytotoxicity (ADCC) and that ADCC likely contributes to back pain syndrome and other muskoloskeletal manifestaions of inflammatory bowel disease [17] we suggested ADCC-driven inflammation as a contributing factor of LBP development. Another potential mechanism is so called “autoimmune pain” caused by IgG-mediated autoimmune response against components of nervous system to which ADCC also contributes [18]. In addition, we demonstrated a pro-inflammatory profile of total plasma N-glycome in chronic LBP patients characterised by the increase in the relative amount of high-branched N-glycan structures, disialylated and trisialylated glycan structures and the decrease of high-mannose glycans and glycans containing bisecting N-acetylglucosamine [19].

Thus, the analysis of glycome may provide important biological and clinically relevant information on the mechanisms of the transition from acute to chronic LBP. Moreover, studies show that changes in glycome profiles may precede the onset of clinical diseases, thus proving glycome as a valuable predictive biomarker. For instance, variation in IgG glycosylation has been shown to precede the onset of rheumatoid arthritis [12,20], type 2 diabetes [21], and hypertension [22].

Herewith, we investigated whether the plasma N-glycome predicts the conversion of acute LBP to chronic LBP using a prospective dataset generated in the framework of the EU FP7 PainOmics project [23,24].

## Material and methods

We prospectively recruited n = 1,114 participants (512 males, 602 females, aged from 16 to 94 with mean age 51.7±15.8) [23,24], through clinical centers across Europe: British School of Osteopathy (under auspicies of King’s College London; KCL, UK), University of Parma (UPR, Italy), San Mateo Hospital (OSM, Italy), St. Catherine Hospital (STCAT, Croatia), and Hospital Oost-Limburg (ZOL, Belgium) (Supplementary table 1). Ethical approval has been obtained at each respective institution, and all participants gave informed consent to take part in the study. Clinical assessment and plasma collection were performed at two time points: at baseline (BL) at the time of their first episode of LBP and follow-up (FU) after three months of conventional treatment. Those who reported ongoing LBP at three months were considered as cases of chronic LBP, while those in whom LBP had resolved were considered controls. Exclusion criteria were: clinically unstable disease, severe psychiatric disorders (apart from mild depression) or mental impairment, recent history (<1 year) of spinal fracture, back pain due to spinal tumour or infection, and pregnancy.

**Table 1.** Comparisons of glycan levels at FU between chronic LBP cases and controls.

| Trait | Current study |  |  | Previous study [19] |  |
| --- | --- | --- | --- | --- | --- |
|  | Effect (SE) | p-value | I<sup>2</sup>, % | Effect (SE) | p-value |
| LB | -0.104 (0.083) | 0.213 | 0 | -0.247 (0.049) | 5.1e-7 |
| HB | 0.108 (0.085) | 0.203 | 0 | 0.223 (0.049) | 6.3e-6 |
| **S0** | **-0.201 (0.089)** | **0.024** | **42.3** | **-0.356 (0.049)** | **4.2e-13** |
| S1 | 0.046 (0.087) | 0.596 | 0 | -0.012 (0.049) | 0.803 |
| S2 | 0.176 (0.091) | 0.054 | 54.6 | 0.254 (0.049) | 2.8e-7 |
| S3 | 0.067 (0.086) | 0.422 | 0 | 0.235 (0.050) | 2.3e-6 |
| S4 | 0.072 (0.086) | 0.405 | 15.8 | 0.057 (0.050) | 0.252 |
| G0 | -0.132 (0.089) | 0.137 | 31.9 | -0.312 (0.044) | 1.9e-12 |
| **G1** | **-0.215 (0.090)** | **0.016** | **41.5** | **-0.330 (0.049)** | **1.6e-11** |
| G2 | 0.132 (0.090) | 0.140 | 60.9 | 0.179 (0.048) | 1.9e-4 |
| G3 | 0.132 (0.086) | 0.125 | 0 | 0.182 (0.048) | 1.7e-4 |
| G4 | -0.017 (0.085) | 0.839 | 0 | 0.128 (0.048) | 7.4e-3 |
| **HM** | **-0.179 (0.087)** | **0.039** | **34.5** | **-0.219 (0.046)** | **1.7e-6** |
| B | -0.149 (0.088) | 0.090 | 0 | -0.281 (0.049) | 8.2e-9 |

Total plasma N-glycans have been assessed at BL and FU as described elsewhere, with 39 original N-glycan traits measured and recalculated to 14 derived glycan traits [19]. At BL, full set of data (both LBP and glycan levels) was present for 1,085 people, while at FU full set of data was available for 549 participants. For 520 participants, information about both LBP and glycan profiles was present at both time points (Supplementary Table 2; Figure 1).

**Table 2.**
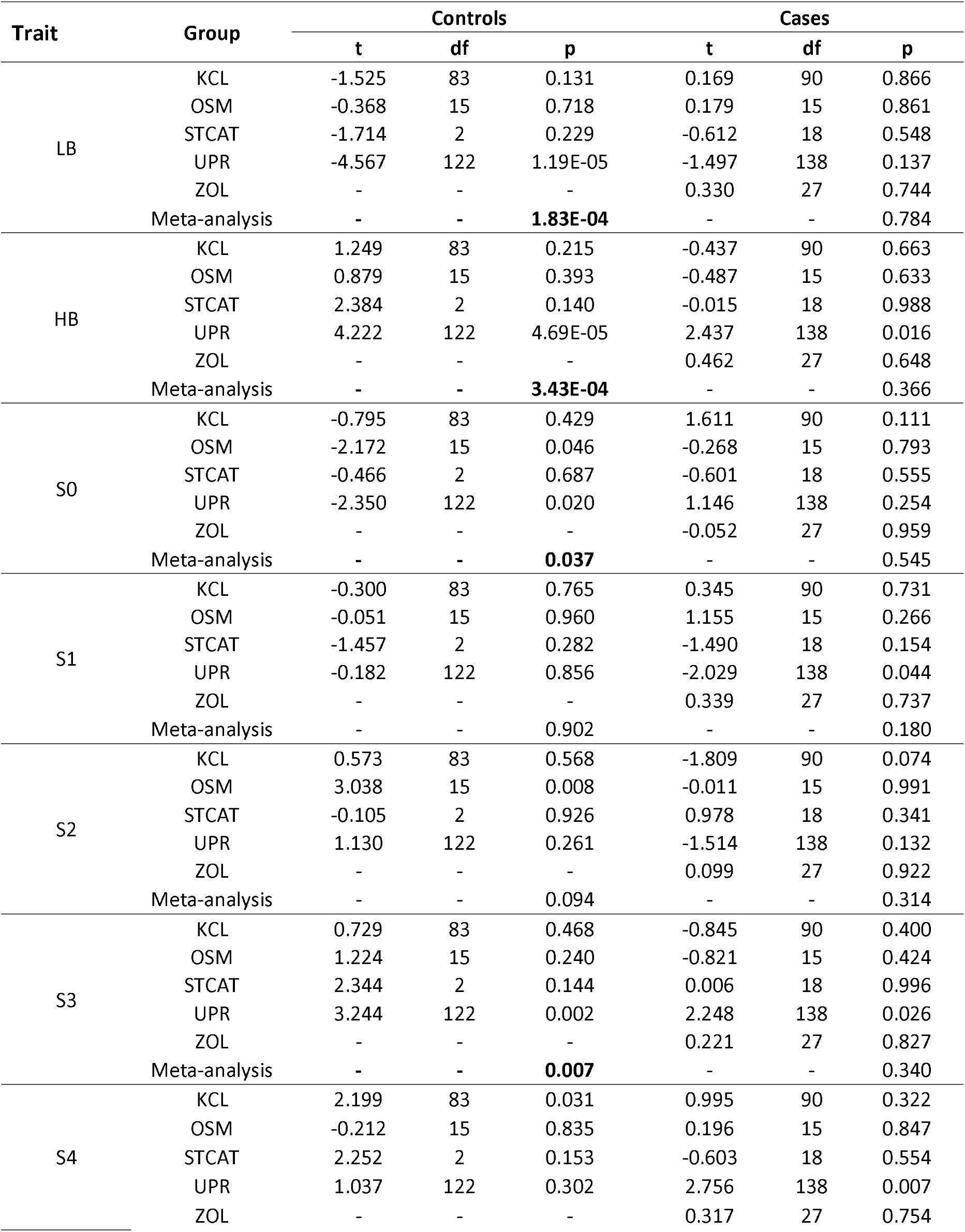
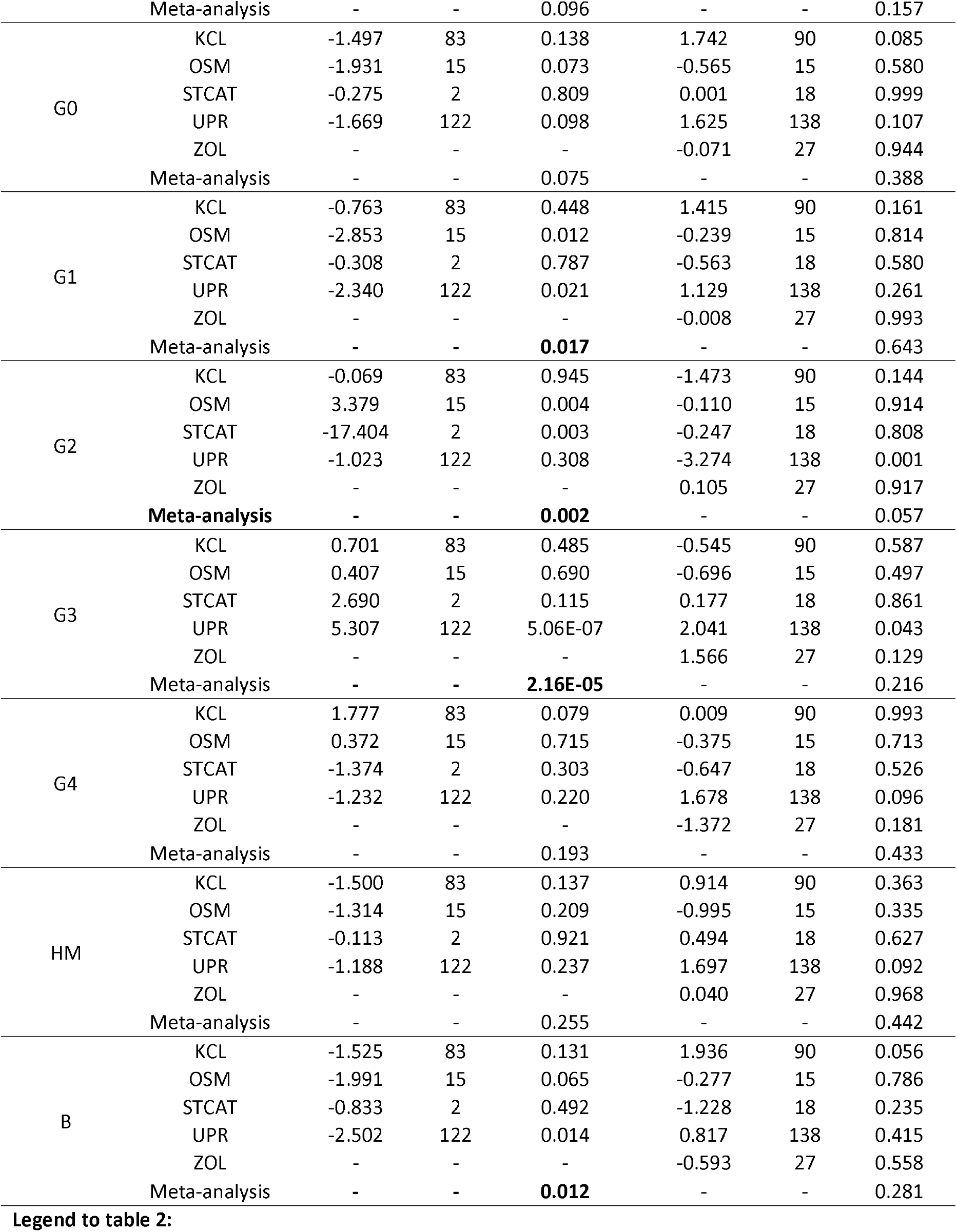
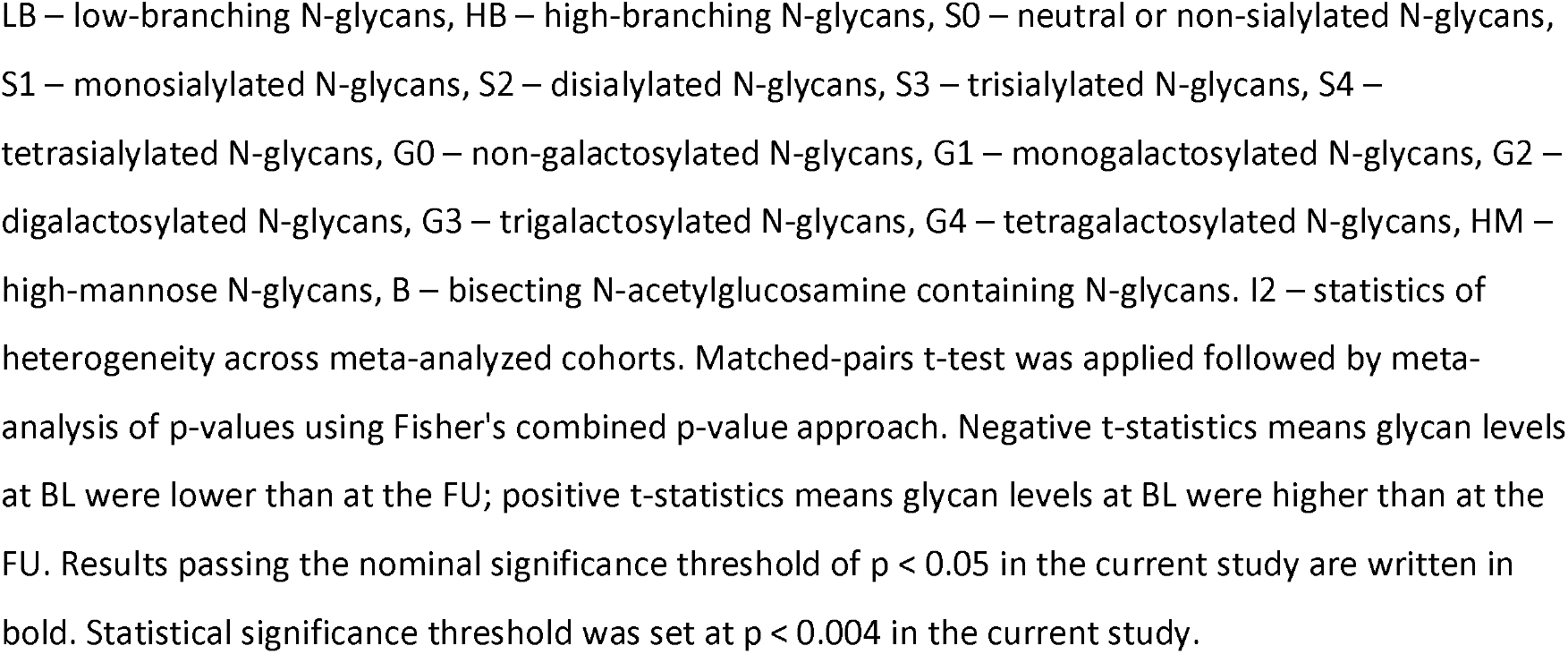
Association between glycan levels in cases of chronic LBP and controlsGlycan trait.

**Figure 1.**
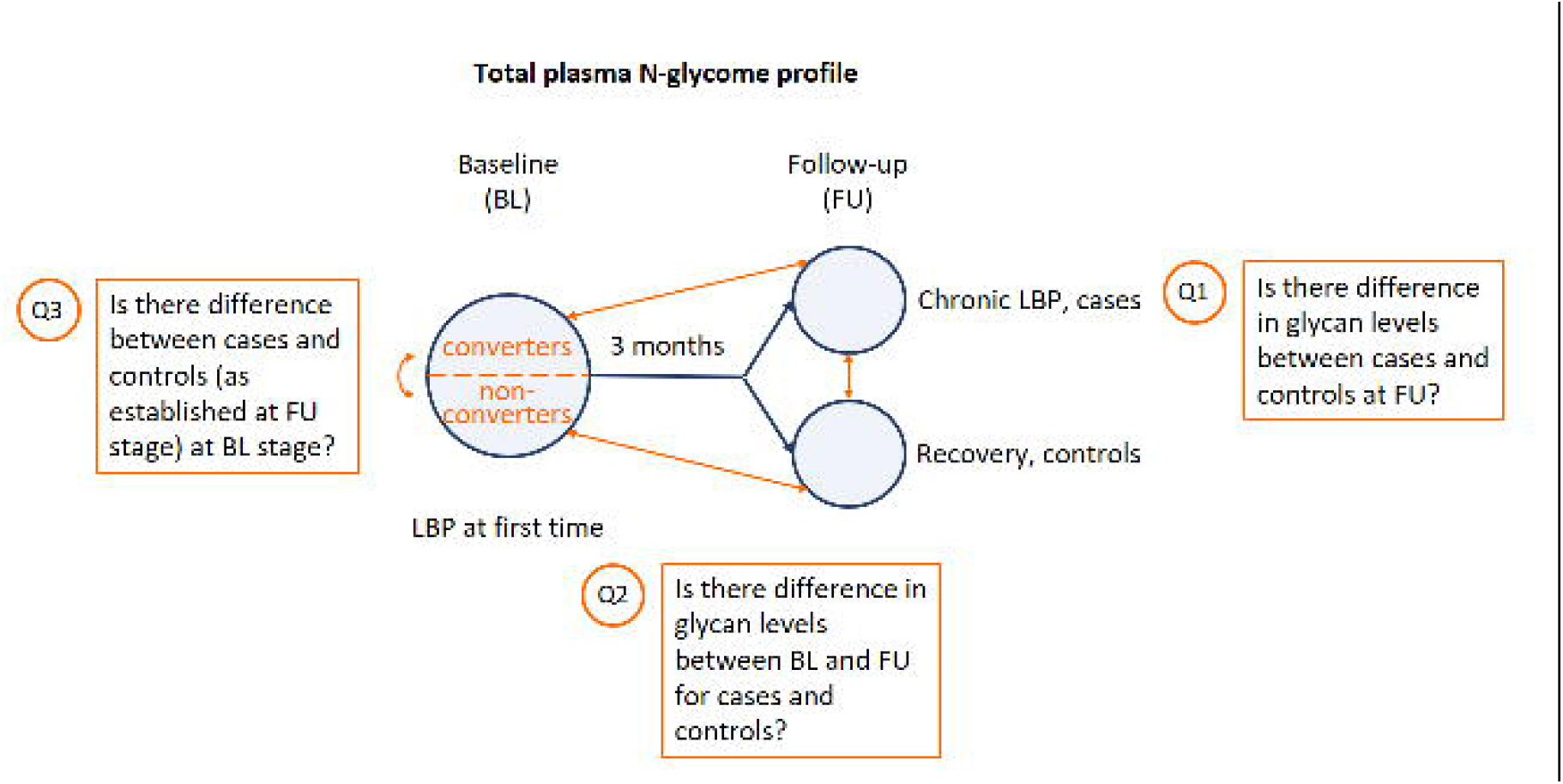
Flowchart of patients’ availability. More details are provided in Supplementary table 2.

We addressed three questions (Figure 2):

**Figure 2.**
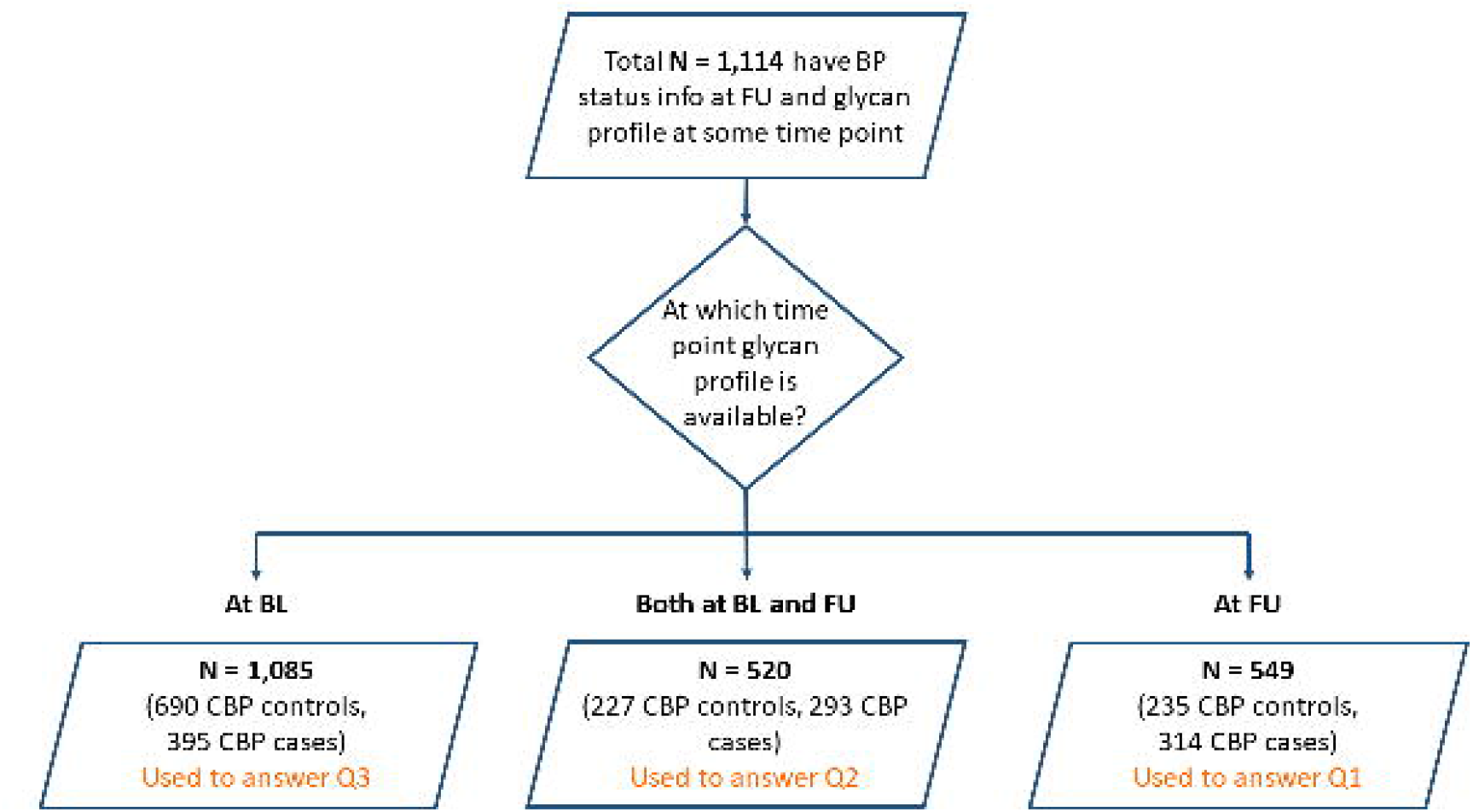
The analysis pipeline. The figure outlines the research questions addressed as well as the design of the corresponding statistical comparisons

- Q1: Is there difference in total plasma N-glycan levels between cases and controls at FU? This is equivalent to a cross-sectional comparison carried out earlier [19] and aimed to assess validity of our dataset;
- Q2: Is there difference in glycan levels between BL and FU for cases and controls? The answer to this question would inform us about dynamics of plasma N-glycome during convalescence and persistence of the disease.
- Q3: Is there difference between cases and controls (as established at FU stage) at BL stage? This would inform us if plasma glycome can serve as predictive biomarker.

For each cohort, glycan profiles were corrected for age, sex, and body mass index via residuals followed by the inverse rank-transformation to normality for the glycan traits to achieve N [0,1]. To address Q1 and Q3 we carried out linear regression considering the case/control status as an explanatory variable and the glycan level as a dependent variable. The results from all cohorts were meta-analyzed with Z-based method using metafor v2.1.0 package for R (https://cran.r-project.org/web/packages/metafor/metafor.pdf). To address Q2, we carried out a paired t-test followed by meta-analysis using metap v1.1 package for R (https://cran.r-project.org/web/packages/metap/index.html). The statistical significance threshold with correction for multiple testing was set at p < 0.05/14≈0.004. All the analyses were run in R programming language v. 3.5.3 and 3.6.1 in RStudio.

## Results

None of the glycan traits was statistically significantly associated with chronic LBP when comparing cases and controls and FU (Q1) and adjusting for multiple testing; however, the direction and the magnitude of the effects were concordant with the results obtained by us previously in a bigger sample [19] (Table 1). Three glycan traits (neutral, S0; monogalactosylated, G1; and high-mannose glycans, HM) achieved nominal significance (p < 0.05).

Addressing Q2, we found statistically significant alteration in plasma glycome between BL and FU in controls, but not in cases (Table 2). The changes concerned levels of low- and high-branching glycans, di- and trigalactosylated glycans (LB, HB, G2, and G3, respectively). Changes with nominal significance (p < 0.05) were identified for neutral (non-sialylated), trisialylated, monogalactosylated glycans, and bisecting glycans (S0, S3, G1, B, respectively). Levels of LB were found to increase at FU, while levels of HB, and G3 decreased at FU compared to BL. Notably, the direction of changes in levels of G2 was not consistent between the cohorts, and most significant results were observed for STCAT with a very small sample size (n = 2); thus, this result must be treated with caution. Considering results of the case-control comparisons at FU (Table 1) and these results, it can be noted that in controls changes are directed towards the plasma N-glycome profile seen in healthy individuals. In particular, LB levels while being decreased in LBP change towards the increase from BL to FU in controls, i.e. individuals whose pain resolved in conventional treatment. At the same time, HB and S3 levels while being increased in LBP change towards the decrease from BL to FU. These findings demonstrate that plasma N-glycome changes along with the process of convalescence towards a “healthy” non-inflammatory profile. In accordance with that, no such changes are seen in cases of chronic LBP, thus assuming the persistence of ongoing inflammation.

Finally, addressing the Q3, we did not find statistically significant differences in plasma N-glycome levels between cases and controls at BL (Table 3).

**Table 3.** Association between glycan levels at BL and chronic LBP.

| Trait | Effect size (SE) | p-value | I<sup>2</sup>, % |
| --- | --- | --- | --- |
| LB | 0.006 (0.066) | 0.930 | 0 |
| HB | -0.002 (0.067) | 0.976 | 0 |
| S0 | 0.021 (0.063) | 0.742 | 25.2 |
| S1 | -0.046 (0.066) | 0.484 | 0 |
| S2 | 0.003 (0.064) | 0.965 | 36.3 |
| S3 | 0.010 (0.067) | 0.881 | 0 |
| S4 | 0.003 (0.065) | 0.966 | 0 |
| G0 | 0.030 (0.064) | 0.636 | 63.2 |
| G1 | -0.013 (0.064) | 0.837 | 20.4 |
| G2 | -0.008 (0.064) | 0.904 | 59.7 |
| G3 | -0.016 (0.067) | 0.814 | 0 |
| G4 | 0.021 (0.066) | 0.750 | 50.7 |
| HM | 0.033 (0.066) | 0.619 | 0 |
| B | 0.020 (0.067) | 0.764 | 31.2 |

## Discussion

The current study aimed to determine whether plasma N-glycome is a predictive biomarker of risk of chronic LBP during an episode of acute severe back pain. We confirmed an earlier observed trend [19] towards the decrease of S0, G1, and HM in patients with chronic LBP, found changes in glycome profile between BL and FU in controls, but not cases of chronic LBP, and did not find differences in glycome profile between cases and controls at BL.

Our findings suggest that plasma N-glycome does not predict transition between acute and chronic LBP; however, it reflects the dynamics of the condition: remains stable if inflammation is progressing and reverses to a normal state during convalescence. This finding corroborates with the results of a recent study of transcriptomic changes in immune cells of those who resolved from LBP after three months of conventional treatment, but not among those who did not resolve [25]. This transcriptomic study identified upregulated neutrophil-driven resolution of inflammation associated with convalescence.

As proven by clinical tools, such as SBT, predicting risks of chronic LBP and disability facilitates treatment and exhibits economic benefits [8-10]. The current study did not provide evidence that total plasma N-glycan levels at LBP onset would be predictive of future chronic LBP, thus warranting to investigate other biomarkers. Among promising options is a polygenic risk score (PRS) that summarises the effects of multiple genetic factors across the genome to predict disease risk [26]. Upon calculation of their PRS, each individual may be classified as high or low risk if their score falls into upper or lower quintile of the PRS distribution. A public resource – PRS atlas – collecting PRS for various diseases has been compiled in response to public and clinical interest in the field [27]. There is now good evidence that this approach works in clinical settings. For instance, PRS added to conventional risk factors could potentially prevent 7% additional cardiovascular events compared to conventional risk prediction [28]. Bronchodilator response to asthma medicine could be predicted by a pharmacogenetic PRS, thereby avoiding unnecessary and potentially dangerous trials of ineffective medicines [29]. PRS predicts more severe cases of inflammatory bowel disease [30]. To the best of our knowledge, apart from our own pilot attempt focusing on glycome-based GWAS [31], no PRS for chronic LBP has been published despite the availability of high-scale GWAS for this trait [32-34], let alone any attempts to combine PRS and clinical tools such as SBT.

Our study limitation is heterogeneity between the study centres. Another limitation is the lack of replication in an independent cohort. However, consistence with the previous study [19] adds confidence in validity of our findings.

## Data Availability

Original data presented in the current study are available through a request to TwinsUK.

https://twinsuk.ac.uk/resources-for-researchers/our-data/

## Acknowledgements

We are grateful to the participants of the study. We also would like to acknowledge Ambreen Tariq, Helena Wells, and Catherine Redshaw for recruiting the study participants as well as the personnel of the following clinical facilities: St Thomas Hospital, Guy’s and St Thomas’ NHS Trust, London; British School of Osteopathy (since 2017, The University College of Osteopathy), London; Darent Valley Hospital, Dartford and Gravesham NHS Trust, Dartford; Northampton General Hospital Trust, Northampton; Solent NHS Trust, Portsmouth; Brighton and Hove City Primary Care Trust (Benfield Valley Healthcare Hub, Brighton Health and Wellbeing Centre, Charter Medical Centre, Sackville Medical Centre); Brockwood Medical Practice, Betchworth; Sea Road Surgery, Bexhill-on-Sea; Woodbridge Hill Surgery, Guildford; St. Catherine Hospital, Zagreb; Clinical Hospital “Holly Spirit”, Special hospital for medicial rehabilitation “Krapinske Toplice”, Medical School University of Split, Clinical Hospital Split; Anesthesia, Critical Care and Pain Medicine Unit, Department of Medicine and Surgery at University of Parma; and Hospital Oost-Limburg, Maastricht.

## Notes

**Funding:** This research was supported by the European Commission FP7 “PainOmics” project (contract No. 602736). YAT and EEE were supported by the Russian Science Foundation grant # 22-15-20037 and the Government of the Novosibirsk region. YSA was partially supported by the budget project of the Institute of Cytology and Genetics # FWNR-2022-0020.

### Competing Interest Statement

YSA and LCK are the founders and owners of PolyOmica and PolyKnomics, private organizations providing services, research, and development in the field of computational and statistical genomics. GL is the founder and owner of Genos Ltd., a private research organization that specializes in high-throughput glycomic analysis and has several patents in this field. IU, ITA, MV, AS, and TP are employees of Genos Ltd. Other authors declare no competing interests.

### Funding Statement

This research was supported by the European Commission FP7 PainOmics project (contract No. 602736). YAT and EEE were supported by the Russian Science Foundation grant # 22-15-20037 and the Government of the Novosibirsk region. YSA was partially supported by the budget project of the Institute of Cytology and Genetics # FWNR-2022-0020.

### Author Declarations

Ethical approval has been obtained at each respective institution who carried out recruitment and sample collection: National Research Ethics Service Committee London - Westminster (London, UK); Comite Medische Ethiek van Ziekenhuis Oost-Limburg (Genk, Belgium); Comitato Etico per Parma (Parma, Italy); IRCCS Fondazione Policlinico San Matteo (Pavia, Italy); Eticki odbor Specijalna bolnica Sv. Katarina (Zagreb, Croatia). Copies of approvals were provided to the European Commission before starting the study. All participants gave informed consent to take part in the study.

